# Intracortical microstimulation of human somatosensory cortex is sufficient to induce perceptual biases

**DOI:** 10.1101/2024.04.19.24305901

**Authors:** Charles M. Greenspon, Natalya D. Shelchkova, Taylor G. Hobbs, Sliman J. Bensmaia, Robert A. Gaunt

## Abstract

Time-order error, a psychophysical phenomenon in which the duration in between successive stimuli alters perception, has been studied for decades by neuroscientists and psychologists. To date, however, the locus of these effects is unknown. We use intracortical microstimulation of somatosensory cortex in humans as a tool to bypass initial stages of processing and restrict the possible locations that signals could be modified. We find that, using both amplitude discrimination and magnitude estimation paradigms, intracortical microstimulation reliably evoked time-order errors across all participants. Points of subjective equality were symmetrically shifted during amplitude discrimination experiments and the intensity of a successive stimulus was perceived as being more intense when compared to single stimulus trials in magnitude estimation experiments. The error was reduced for both paradigms at longer inter-stimulus intervals. These results show that direct activation of primary somatosensory cortex is sufficient to induce time-order errors.

## Introduction

Psychophysical measures of the limits of human perception were some of the earliest forays into understanding how the brain processes sensory information. One of the earliest acknowledged and most studied psychophysical phenomena is that of the time-order errors (TOEs): where an interaction occurs between two sequentially presented stimuli leading to a perceptual bias of either stimulus (1–4). Indeed, these findings have shaped experimental design for the last century as scientists endeavor to minimize confounds when presenting many successive stimuli. The most common observation with pairs of stimuli is that the second stimulus tends to be perceived more intensely than expected, typically referred to as an enhancement effect (5–10). Research has attributed these sequential order effects to memory (6, 11), attention (5, 12), and gain modulation (10, 13, 14). Consistent with these hypotheses, the magnitude of the error has been shown to be in part dependent upon the inter-stimulus interval (ISI) (15). While all of these hypotheses indicate top-down modulation, the locus through which this is affected is unclear. Alternatively, modulation of tactile sensations has been shown to occur in both the brainstem (16, 17) and in the thalamus (18, 19), suggesting that time-order errors could also occur as a result of sub-cortical regulation.

Intracortical microstimulation (ICMS) provides a novel avenue for studying psychophysical phenomena (including time-order errors) as it allows for direct stimulation of cortical regions, thus bypassing lower order structures and mitigating any effects they may have on perception. Consequently, ICMS allows us to probe higher-level structures in the absence of bottom-up effects such as adaptation of the peripheral afferents (20). Additionally, examining these phenomena with ICMS allows us to question the extent to which ICMS mimics natural tactile stimulation. If ICMS evokes similar phenomena, then we can deduce that it is recruiting neural structures with reasonable similarity to natural sensation. Alternatively, if ICMS is not susceptible to these psychophysical effects, experimental design will need to be adjusted accordingly. The implications for experimental design are of particular importance as brain computer interfaces that include ICMS become more common (21–24), and researchers seek to fully understand their findings and optimize their devices appropriately.

In this study, we provide ICMS to three participants with cervical spinal cord injuries with microelectrode arrays implanted into Brodmann’s Area 1 (A1) of somatosensory cortex. This robustly evokes tactile sensations on their contralateral hand while manipulation of the stimulus amplitude modulates the perceived intensity of the sensation (21, 25, 26). We implemented both amplitude discrimination and magnitude estimation paradigms (3) to probe the perceptual consequences of sequential stimulation with a view of understanding whether time-order effects can be entirely attributed to higher-order cortical regions or if, instead, preceding neural structures (mechanoreceptors, dorsal root ganglia, cuneate, or thalamus) play a significant role in these perceptual biases. Our investigation revealed that time-order errors arose when using ICMS with both paradigms and that the magnitude of the error was influenced by the inter-stimulus interval.

## Results

### Interval biases are revealed by amplitude discrimination

Time-order errors often present as recency bias: when observed, stimuli that occur closer to the time at which the participant makes a judgement are reported as feeling more intense than those that occurred earlier (6). In amplitude discrimination experiments, where a participant is asked to compare the intensity of two stimuli presented in succession, this results in a bias towards reporting the second stimulus as more intense. To test whether this occurs with ICMS, we implemented a 2-alternative forced choice (2AFC) amplitude discrimination task (**Figure 1A**). Briefly, the participant was cued using a fixation cross presented on a computer monitor and two ICMS trains were delivered during which the color of the fixation cross was changed to green. The participant was asked to report which of the two stimuli was more intense. On each trial, a standard stimulus train of 60 μA and a comparison stimulus train between 40 and 80 μA were given (all stimulus trains were 1 second long, 50 Hz, and separated by a 1 second ISI). Importantly, trials were counterbalanced such that there was an equal proportion of trials in which the standard stimulus train was delivered in the first or second interval and all unique stimulus pairs were delivered in a block format where stimulus order was randomized for each block. The participants were asked to report which interval contained the stronger stimulus train and the proportion of trials in which the interval containing the comparison stimulus train was reported as being stronger was computed (**Figure 1B, C**).

**Figure 1.**
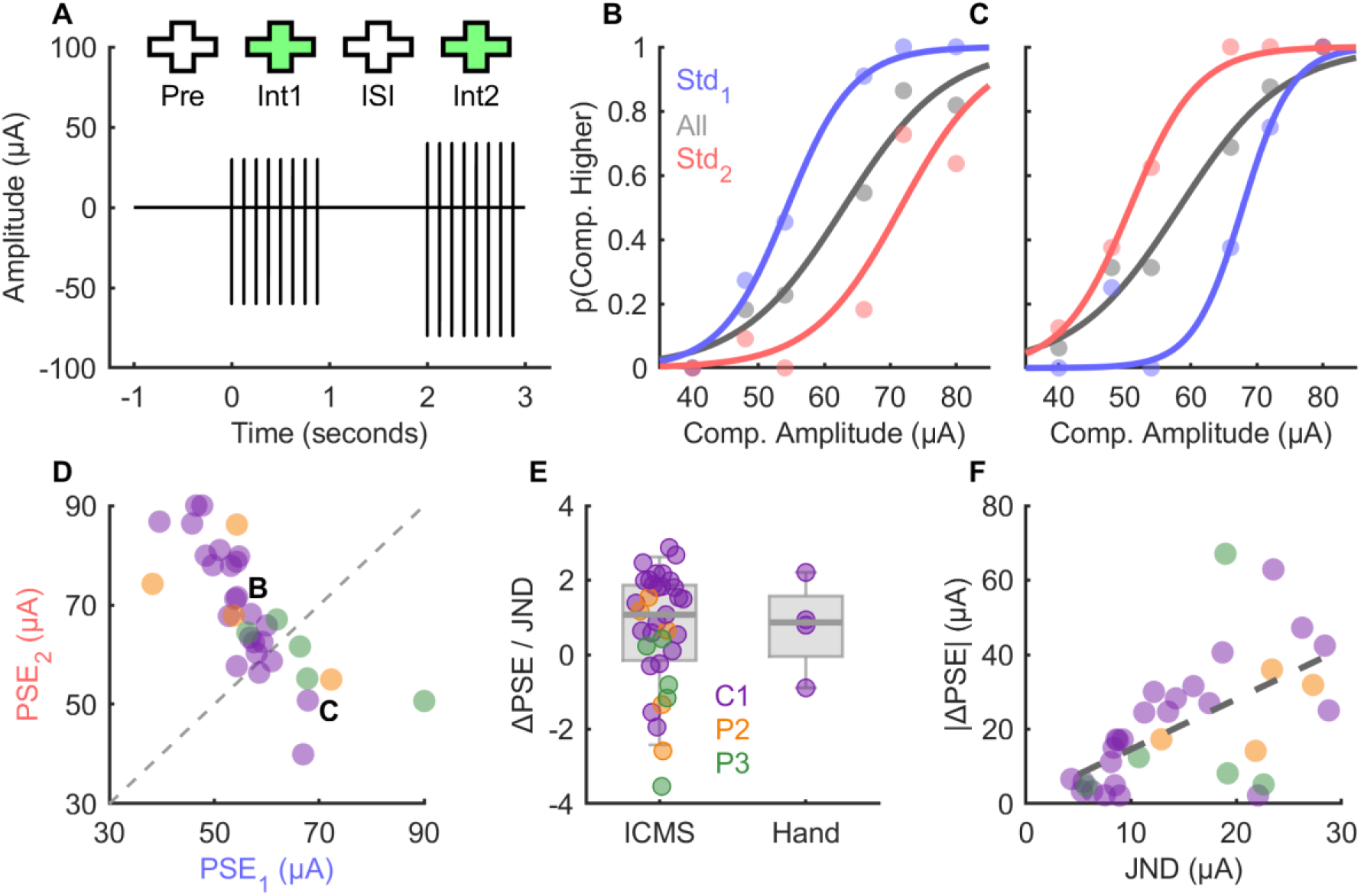
Amplitude discrimination reveals order dependent psychometric functions. **A)** Task schematic for an amplitude discrimination trial. Two ICMS trains are delivered, the participant is cued with a fixation cross, and the participant is asked to report which of the two stimuli was more intense. Note that the cathodic (negative) peak indicates train amplitude. **B,C)** Example psychometric functions for two electrodes when trials are split into those in which the standard stimulus (60 μA) was presented in the 1^st^ or 2^nd^ interval, or when they are combined. Separation of these curves indicates order-dependent biases. **D)** PSEs with respect to standard interval across all electrodes. Labels indicate example electrodes shown in panels **B & C**. PSE outliers are capped at 30 or 90 μA for illustrative purposes. **E)** The normalized order effect for both ICMS and tactile amplitude discrimination tasks. **F)** Amplitude sensitivity (JND) of each electrode and the absolute difference in the PSE.

To measure participant performance for each electrode, a sigmoid function was fit to each of these points. From this fit, the point of subjective equality (PSE: the point at which the sigmoid equals 0.5) and the just noticeable difference (JND: the difference in amplitude required for the participant discriminate the comparison stimulus from the standard stimulus on 75% of trials) were derived. Participants were able to perform this task across the vast majority of electrodes tested (**Supplementary Figure 1A**), however the results were discarded for the two electrodes that they could not use for the task (n = 2).

After splitting the trials into those where the standard stimulus was presented in the first or second interval (**Figure 1B, C**) we compared their psychometric fits. As indicated from the example psychometric plots shown, we found a systematic effect of stimulus order upon the PSE (**Figure 1D**, Wilcoxon rank sum test, Z_[33]_ = 4.10, p = 0.8e^-4^). Participants would consistently rank one interval as being more intense than the other resulting in differences in the PSEs (**Supplementary Figure 1B**, adjusted coefficient of determination, R^2^ = 0.65, F_[33,31]_ = 17, p = 0.9e^-5^). This effect occurred for each participant on a subset of electrodes, though to varying degrees (permutation test p < 0.05: 14/25 for C1, 3/5 for P2, and 1/5 for P3). The majority of electrodes reveal a systematic bias where the comparison stimulus was perceived as being more intense when it was in the second interval (**Figure 1D**, points above and to the left of the unity line; example in **Figure 1B**), consistent with tactile literature on enhancement (10); however, it should be noted that at least one electrode from each participant showed the opposite effect (**Figure 1D**, points below and to the right of the unity line; example in **Figure 1C**). Crucially, while the plots show when the comparison was enhanced in the second interval, the standard stimulus was also susceptible to enhancement when in the second interval. Thus, participants also systematically underestimated the intensity of the comparison stimuli in the first interval, resulting in symmetrical shifts from the mean.

Next, we the investigated it stimulus order had an effect on the discrimination performance (JND) across electrodes but found no effect (**Supplementary Figure 1C**, Wilcoxon rank sum test, Z_[33]_ = 2.02, p = 0.08 after Bonferroni correction), though there was a tendency for the JNDs to be higher when the standard was in the second interval. Finally, to contextualize the effect, we implemented a similar paradigm with mechanical stimulation of one participant’s ipsilateral hand (C1 has normal sensation in the ipsilateral hand). After normalizing the effect size by the sensitivity to standardize units (**Figure 1E**), we observed that the total magnitude of the effect was similar across stimulus methods (Wilcoxon rank sum test, Z_[33,4]_ = 0.025, p = 0.98), and with equal variation (2-sample F-test, F_[32,3]_ = 1.29, p = 0.967; permutation test, p = 0.98).

We then sought to find an explanation for the diversity in the effect across electrodes. We observed that there was a moderate relationship between the absolute ΔPSE and the JND (**Figure 1F**, adjusted coefficient of determination, R^2^ = 0.33, F_[33,31]_ = 17, p = 0.25e^-3^) suggesting that the effect was magnified for less sensitive electrodes, consistent with the idea that there is an underlying bias that can be overpowered with sufficient information (i.e., amplitude-sensitive electrodes). We also examined if the detection threshold of the electrode might influence the ΔPSE but found no effect (adjusted coefficient of determination, R^2^ = 0.05, F_[33,31]_ = 2.7, p = 0.11). Finally, we repeated the experiment with multiple frequencies interleaved (50, 100, and 200 Hz) on a subset of electrodes (n = 5) and examined if the ΔPSE or ΔJND systematically varied with frequency. We found substantial but not systematic variation on the ΔPSE (**Supplementary Figure 1D**, 2-way ANOVA_[ΔPSE, frequency]_, F_[2,17]_ = 1.18, p = 0.35) whereas there was a weak but significant decrease in the ΔJND as frequency increased (**Supplementary Figure 1E**, 2-way ANOVA_[ΔJND, frequency]_, F_[2,17]_ = 4.13, p = 0.0492), though this is likely a result of the general tendency for JNDs to decrease with frequency (**Supplementary Figure 1F**, 2-way ANOVA_[JND, frequency]_, F_[2,17]_ = 7.6, p = 0.0098).

### Sequential magnitude estimation reveals an enhancement effect

Amplitude discrimination paradigms require that the participants hold both sensations in memory and are therefore susceptible to memory effects and recency biases (6). To determine if the observed effect should be attributed to a memory effect or an enhancement effect, where the preceding stimulus causes the subsequent stimulus to feel more intense (10), we implemented a sequential magnitude estimation paradigm. Much like the amplitude discrimination task (**Figure 1A**), two stimuli (1 second duration, 50 Hz) were delivered with a 1 second ISI and were cued with fixation crosses. However, in this experiment the participant (C1) was instructed to only report the intensity of the second stimulus (cued by the second green fixation cross). This paradigm removes any memory effects and allows probing of the subjective intensity of the most recent stimulus without any attentional effect as the participant always knew when and where to expect the second stimulus.

Consequently, we compared the reported intensity of the second stimulus when there was a conditioning stimulus (on the same electrode between 10 and 80 μA) or not (catch) and found enhancement of the second stimulus when a conditioning stimulus was present (example shown in **Figure 2A**). Furthermore, the magnitude of this effect was determined in part by the strength of the conditioning stimulus (**Figure 2B**) where more intense conditioning stimuli resulted in more enhancement (3-way ANOVA [conditioning amplitude, ISI, test amplitude], F_[3, 1193]_ = 24.9, p < 0.01). Next, we repeated the paradigm but kept the intensity of the conditioning stimulus equal and instead varied its duration. We found, in keeping with the prior result, that longer conditioning stimuli tended to induce more enhancement (**Figure 2C**, 3-way ANOVA [conditioning duration, ISI, test amplitude], F_[3, 972]_ = 16.66, p < 0.01). Finally, we compared whether the location of the conditioning electrode played a role in the enhancement by providing a 80 μA conditioning stimulus to an adjacent electrode with a nearby projected field (interleaved with catch trials and conditioning stimuli on the same electrode) and found that the effect was reduced for all tested electrodes but was not completely abolished for two of the five (**Figure 2D**, 2-way ANOVA _[conditioning location, test amplitude]_, F_[1, 598]_ = 13.1, p < 0.01; note that the ANOVA was constructed as same/different conditioning electrode instead of electrode number). These results indicate that the order effect observed with the amplitude discrimination experiments is likely a byproduct of the enhancement effect observed here.

**Figure 2.**
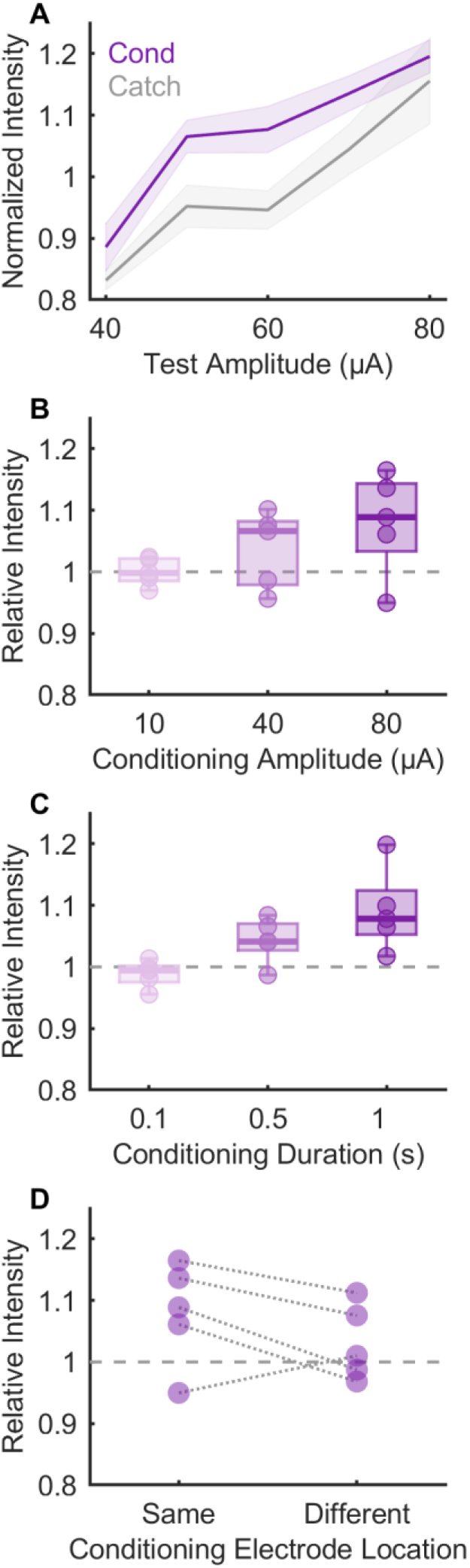
Sequential magnitude estimation shows potentiating effects. **A)** Example normalized intensity reports in the presence (cond) or absence (catch) of an 80 μA conditioning stimulus. Mean and SEM shown. **B)** Mean test intensities for each electrode relative to the catch intensity for different conditioning amplitudes (all 1 s in duration) or **C)** durations (all at 80 μA). **D)** Relative intensity for each electrode when the conditioning stimulus was on the same or an adjacent electrode. All data from C1.

### Enhancement effects decay with inter-stimulus interval

Interactions between successive stimuli leading towards increases in percept intensity depends on the ISI (14, 27). A potential explanation for this phenomenon is that the brain continuously integrates information within a finite window, and at greater ISIs the initial stimulus falls outside that integration window. To test whether ICMS is consistent with a cortical integration mechanism, we performed both amplitude discrimination and magnitude estimation experiments with either 1 or 5 second ISIs (and 6 second inter-trial intervals). When comparing psychometric functions across the two conditions (**Figure 3A inset**), order effects were significantly reduced when the ISI was 5 seconds (**Figure 3A**, Wilcoxon rank sum test, Z_[33]_ = 3.37, p < 0.01). Crucially, this was true for both positive and negative ΔPSEs, with the distribution of the ΔPSE at 5 seconds being much smaller than those at 1 second (2-sample F-test: F_[32,32]_ = 7.4, p < 0.01). Furthermore, the sequential magnitude estimation paradigm revealed a similar result (**Figure 3B,C**) where ISI had a significant effect on the perceived intensity of the test stimulus (4-way ANOVA _[electrode, conditioning amplitude, ISI, test amplitude]_: F_[1, 1745]_ = 99.32, p < 0.01). Finally, we repeated the amplitude discrimination experiments on a subset of electrodes with 4 inter-stimulus intervals (0.5 – 5 seconds, **Figure 3D**) and found a similar trend, though more variation was observed at the shortest ISI than might be expected (**Figure 3E**).

**Figure 3.**
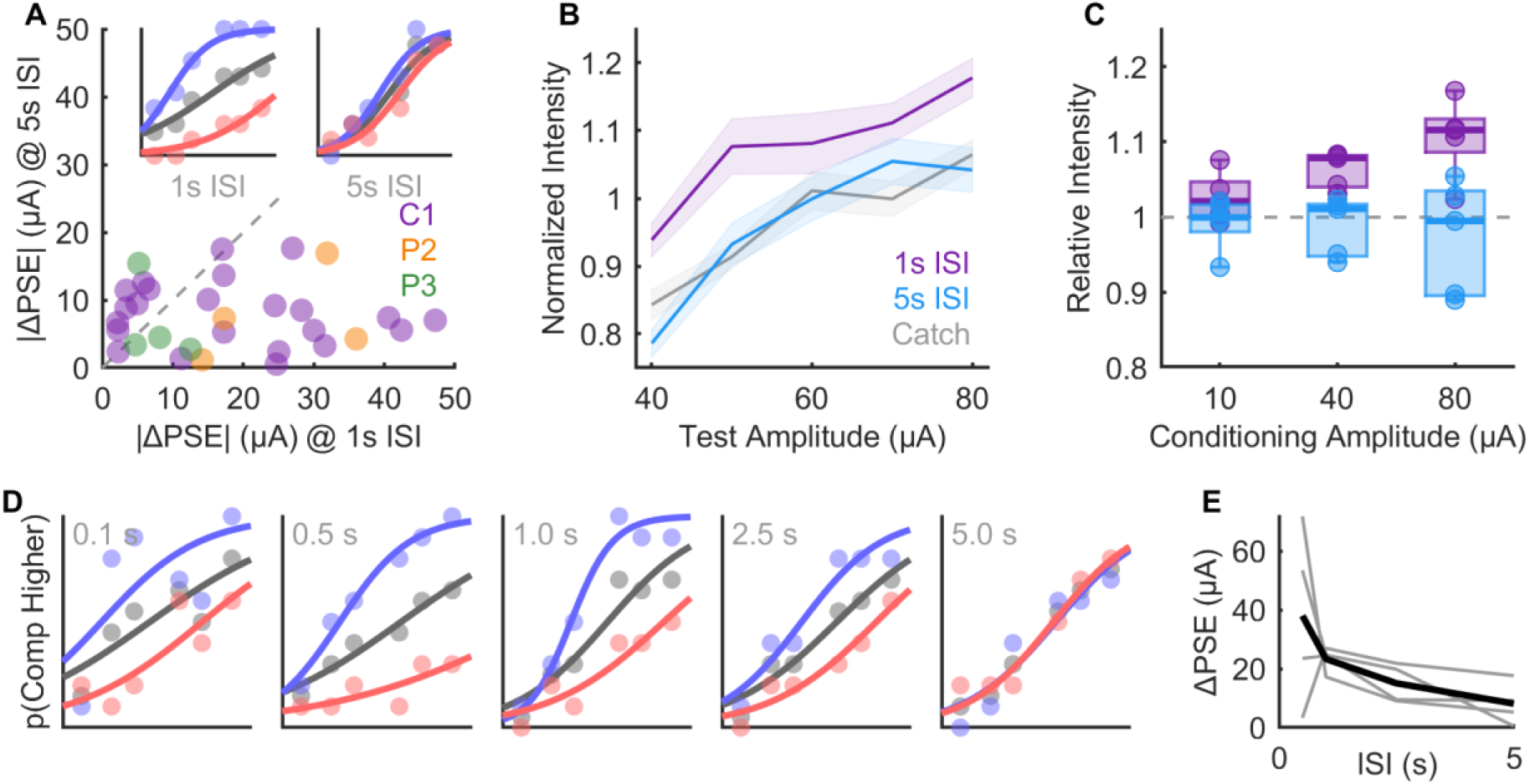
Sequential effects are ISI dependent. **A)** The absolute ΔPSE when amplitude discrimination is performed with a 1 or 5 second ISI. **Inset)** Example psychometric functions for an example electrode at each ISI. **B)** Example normalized intensity reports for an example electrode when sequential stimuli are delivered with either a 1 or 5 second ISI. Catch stimuli indicate when the first interval contained no stimulus. **C)** Test intensities at both ISIs relative to the catch stimulus for different conditioning amplitudes (all 1 second in duration). **D)** Psychometric functions from a single electrode at 5-different ISIs. **E)** Absolute ΔPSE from 4 electrodes at multiple ISIs. Data in panels B-E from C1.

## Discussion

### Enhancement dependent order effects are mediated by higher order structures

Intracortical microstimulation allows for precise stimulation of cortical structures; this bypasses any modulation or gating that may occur at earlier structures (mechanoreceptors, dorsal root ganglia, brainstem nuclei, or thalamus). It does not, however, prevent higher order structures such as secondary somatosensory cortex or prefrontal cortex from exerting top-down control upon S1. The fact that time-order errors were observed in all 3 participants tested (**Figure 1D**) reveals that lower order structures are not necessary for these phenomena. These results do not, however, preclude the possibility that these effects are mediated by bottom-up effects that start in Area 1, potentially including thalamocortical synchronization (28), local rebound excitation (29), or spatial attention (30). Similar psychophysical phenomena such as adaptation (20, 31–33) are also likely mediated by higher order structures, though further testing is necessary to confirm this.

### Adaptation and enhancement occur over different timescales

The results shown here imply that over certain timescales, stimuli should get more and more intense (**Figure 1**, **Figure 2**). However, existing vibrotactile (20, 32, 34), peripheral nerve stimulation(33), and even other ICMS (35) literature implies that repeated stimulation should desensitize participants. Though these results may seem in conflict, they in fact occur together. Indeed, in magnitude estimation experiments we typically normalize all ratings within blocks to offset a global reduction in reported intensities caused by desensitization, though the degree to which this occurs is electrode dependent. Short term enhancement effects thus occur relative to the level of global sensitivity on each trial.

### ICMS activates neural structures in a naturalistic manner

The fact that the magnitude of the observed TOE effects evoked by ICMS were similar to those driven by natural touch (**Figure 1E**) suggests that the activity of higher-order cortical areas in response to ICMS is sufficiently similar to what occurs naturally. The fact that the degree of TOE observed was determined in part by the intensity or duration (**Figure 2B,C**) of the first stimulus (7–9) and the inter-stimulus interval (**Figure 3**) (15), in keeping with existing literature, implies that the neural mechanisms are similar between ICMS and touch. The one caveat we observed, however, is that at very short ISIs, the relationship became much less consistent (**Figure 3E**). We posit that this is due to the kind of idiosyncratic rebound excitation or inhibition observed and simulated with ICMS immediately following a stimulation train (29, 36).

### Implications for BCI experiments

These findings further cement the notion that ICMS reliably activates neural structures in naturalistic ways and is susceptible to the same effects observed with natural touch (and likely other natural stimuli). Furthermore, these results imply that there are likely to be well established phenomena in the psychophysical literature that risk confounding ICMS experiments and researchers should take this into consideration when designing experiments. In particular, amplitude discrimination experiments, a pillar of ICMS experiments for determining electrode sensitivity, must either include long ISIs, or be explicitly counterbalanced to ensure equal stimulus presentation weighting. Furthermore, magnitude estimates should be performed using long inter-trial intervals (>3 seconds) to minimize the interactions between successive stimuli.

## Data Availability

All data is available at the Data Archive for the BRAIN Initiative.

https://dabi.loni.usc.edu/dsi/UH3NS107714

## Acknowledgments

We would like to thank the participants for their generous contribution to the advancement of science. Additionally, this work would not have been possible without the teams that compose the Cortical Bionics Research Group at the universities of Pittsburgh, Chicago, and Northwestern. The work at the University of Chicago and University of Pittsburgh was supported by NINDS grants UH3 NS107714 and R35 NS122333.

## Methods

### Participants

This study was conducted under an Investigational Device Exemption from the U.S. Food and Drug Administration and approved by the Institutional Review Boards at the Universities of Pittsburgh and Chicago. The clinical trial is registered at ClinicalTrials.gov (NCT01894802). Informed consent was obtained from all participants before any study procedures were conducted. Participant C1 (m), 55-60 years old at the time of implant, presented with a C4-level ASIA D spinal cord injury (SCI) that occurred 35 years prior to implant. Participant P2 (m), 25-30 years old at the time of implant, presented with a C5 motor/C6 sensory ASIA B SCI that occurred 10 years prior to implant. Participant P3 (m), 25-30 years old at the time of implant, presented with a C6 ASIA B SCI that occurred 12 years prior to implant.

### Cortical implants and stimulation

Four microelectrode arrays (Blackrock Neurotech, Salt Lake City, UT, USA) were implanted in each participant. Two arrays were implanted in the hand representation of Brodmann’s area 1 of somatosensory cortex and two arrays were implanted in the hand and arm region of motor cortex. The two arrays in BA1 were 2.4 mm x 4 mm, composed of 60 electrodes 1.5 mm in length, wired in a checkerboard pattern such that 32 electrodes could be stimulated. Two percutaneous connectors (Blackrock Neurotech) were fixed to the skull, each attached to one motor and one sensory array. ICMS was delivered through each electrode with a CereStim 96 (Blackrock Neurotech) stimulator. The stimulus pulses consisted of a 200 μs cathodic phase followed by a half-amplitude 400 μs anodic phase (to maintain charge balance), with a 100 μs interphase duration.

### Amplitude discrimination

All amplitude discrimination experiments were conducted on electrodes with detection thresholds below 40 μA. For each trial a 60 μA standard was used and 6 comparison stimuli ranged between 40 and 80 μA (excluding 60 μA). Participants were cued to the stimuli with a fixation cross. One second before trial start a white fixation cross would appear to indicate that the trial was about to begin, it would turn green when the first stimulus was presented, revert to white for the inter-stimulus interval, and then turn green again during the second stimulus interval. For each electrode the trials were counterbalanced, randomized, and presented in a block format (minimum of 8 blocks), totaling at least 192 trials (6 amplitudes, 2 orders, 2 ISI, 8 blocks). An inter-trial interval of 5 seconds was used for all experiments not including the duration for participants to respond. Participants were encouraged to discard trials that they were either unsure of or had not paid attention to for whatever reason to minimize uncertainty biases. Any discarded trials were repeated at the end of the block. Most experiments used 50 Hz stimulation though a subset used 100 or 200 Hz stimulation and are described as such in the main text. Data were collected from 25 electrodes in participant C1, and 5 electrodes in both participants P2 and P3. For the mechanical version of the task, a linear stage (V308, Physik Instrumente, Germany) was used to control a 5 mm diameter aluminum probe with a beveled tip. Stimuli were trapezoidal in shape, with the standard stimulus being 1 mm and the comparison stimuli being between 0.85 and 1.15 mm in depth. To conserve the profile of the trapezoid, speeds varied (4.25 – 5.75 mm/s) such that the transient components of the trapezoid were 0.1 seconds and the stationary phase was 0.8 seconds (totaling 1 second duration).

To produce psychometric functions, the proportion of trials in which each comparison stimulus was reported as being more intense was computed. Then, least-squares optimization was used to fit a 2-term logistic function to these points. Both the x-offset and slope of the function were left unconstrained. The JND was computed as half of the difference of the inverse of the sigmoid where *p* equaled 0.25 and 0.75. To determine the ΔPSE between orders, the above fits were repeated after the trials were split according to the interval in which the comparison stimulus was presented. The ΔPSE was thus computed as the difference in the x-offset between the two functions. As the ΔPSE is equivalent to the interval bias (**Supplementary Figure 1B**), individual electrode effects were tested for significance with a binomial test.

### Magnitude estimation

Magnitude estimation experiments were only performed in participant C1. Each trial followed a similar format to the amplitude discrimination experiments regarding fixation crosses, randomization, block design, inter-trial intervals, and discard strategy but did not require counterbalancing. Participant C1 was explicitly told to ignore the first stimulus and only consider the intensity of the stimulus presented in the second interval for all reports. On a subset of trials, no stimulus was given during the first interval but the participant was still cued to the interval – these are termed catch trials. Again, trials with 1 or 5 second inter-stimulus intervals were fully interleaved. On the small percentage of trials where the participant did not detect any stimulus in the second interval the trial was discarded. All magnitude estimation experiments used 50 Hz stimulation. The normalized intensity was computed for each electrode on a per-block basis by dividing the reported intensity on each trial to the mean intensity across the entire block.

To compute the relative effect of the conditioning stimulus, the reported magnitudes for each treatment condition at each amplitude were expressed as a fraction of the mean of the catch intensity at each test amplitude. This allowed for within amplitude normalization before values could be combined across amplitudes. Statistical effects were assessed using N-way ANOVA (*anovan*, Matlab) with the conditioning duration/amplitude, ISI, test amplitude, and test electrode provided as appropriate.

## Data & Code Availability

All data is stored at the Data Archive BRAIN Initiative (https://dabi.loni.usc.edu/dsi/UH3NS107714) and code for analysis is available upon request.

## Supplementary Figures

**Supplementary Figure 1.**
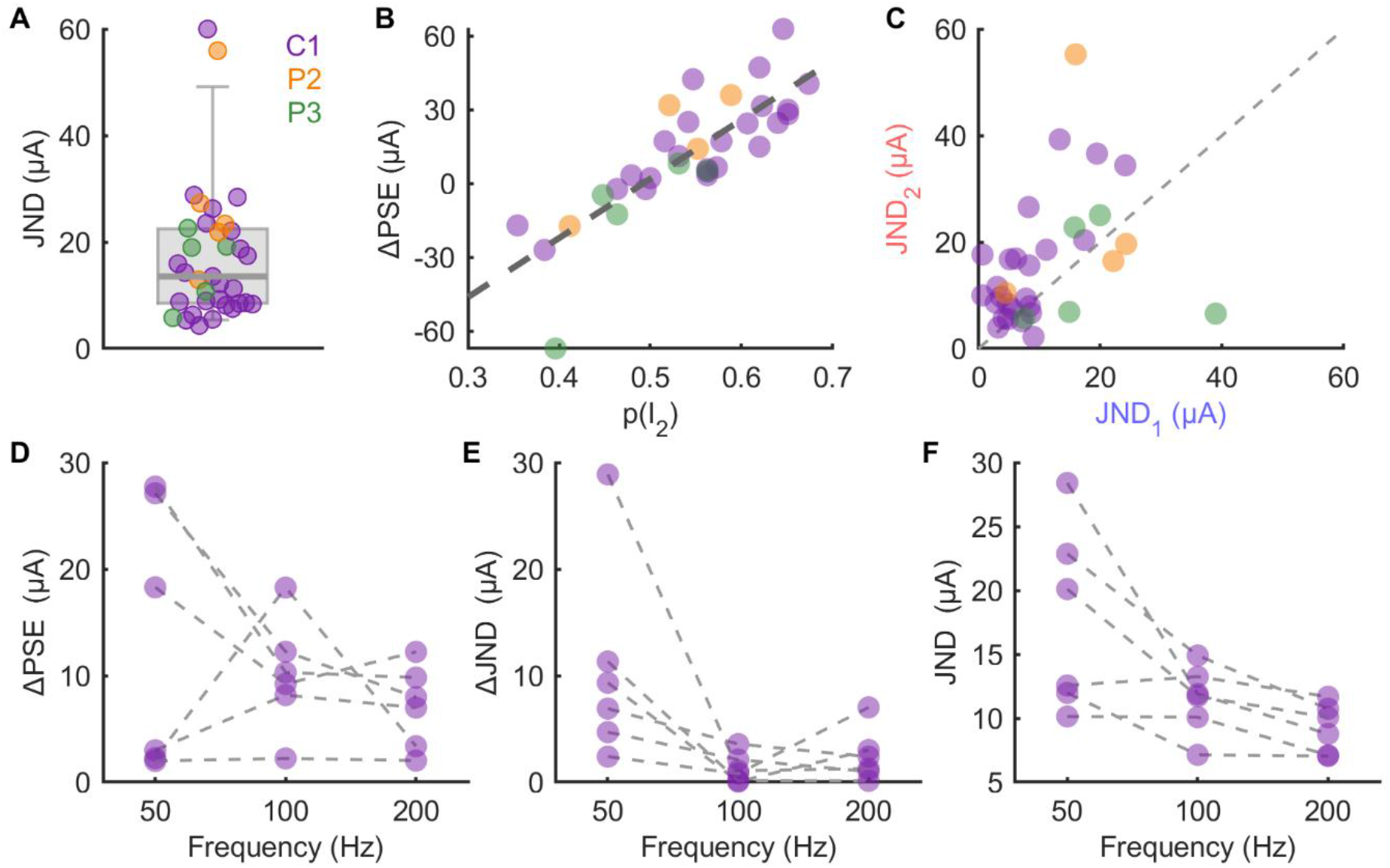
**A)** JNDs for all electrodes tested with a 1s ISI. Box and whiskers indicate 5^th^, 25^th^, 50^th^, 75^th^, and 95^th^ percentiles. **B)** The relationship between the proportion of trials in which the participant selected the second interval and the computed ΔPSE. **C)** JND computed on the subset of trials in which the standard was in the first interval (abscissa) versus those in which the standard was in the second interval (ordinate). **D)** The effect of frequency on ΔPSE, **E)** ΔJND, and **F)** JND.

